# Distribution of Obesity-related Health Outcomes Across the Urban-Rural Commuting Area in Mississippi, Alabama, Louisiana, and Georgia

**DOI:** 10.1101/2023.02.04.23285474

**Authors:** Roungu Ahmmad, Fazlay Faruque

## Abstract

Obesity-related chronic diseases are still major public health concerns in the United States, particularly in the south. The purpose of this study is to examine the association between obesity-related chronic diseases and rurality/urbanicity in the four Deep South states-Mississippi, Alabama, Louisiana and Georgia. We used publicly available Zip Code level approximations of USDA-developed RUCA Code for rurality designation and Zip Code level PLACES data developed by CDC for selected obesity-related health outcomes-Asthma, Obesity, Chronic Obstructive Pulmonary Disease, Coronary Heart Disease, Diabetes, High Cholesterol, Stroke and High Blood Pressure. This study employed the random forest method, partial least squares discriminant analysis and multinomial logistic regression to investigate the association between selected health outcomes and degrees of rurality. There are significant differences in the prevalence of Asthma, Obesity, COPD and Stroke between Metropolitan and small towns or complete rural areas. On the other hand, while considering Micropolitan and small towns or complete rural areas, Asthma, Obesity, COPD, Diabetes, High Cholesterol and Stroke show significant differences in prevalence. This study revealed disparities in health outcomes per RUCA Codes, which can be useful to target specific geographic areas for appropriate interventions.

## 1. Introduction

In the United States, obesity is increasingly causing major health issues that impact the country’s economy, health, and even military readiness [1]. Over one-fourth of all children and, more than one-third of all adults in the US are obese, according to the Centers for Disease Control (CDC) [2]. CDC also mentions that more than half of Americans do not live within half a mile of a park, and 40% of all US households do not live within one mile of healthier food retailers [2]. The situation does not seem to be improving, and by 2030, almost half of adults in the US are projected to be obese [3]. Southern states are burdened more with obesity-related public health issues than other parts of the US [4]. MS, AL, and LA are consistently among the topmost obese states; currently, MS is number one, AL is third, and LA is fourth in the ranking [4]. Among the so-called Deep South states, only South Carolina (SC) and GA are in relatively better shape. In this study, we included three states, MS, AL, and LA, which are among the top five obese states. Among these four contiguous states, GA has better health status than the other three. In the US, the overall adult obesity prevalence percent in 2020 is lowest in Colorado (24.2) and it is highest in MS (39.7), which clearly stands out in the standard deviation map shown in Figure 1.

**Figure 1.**
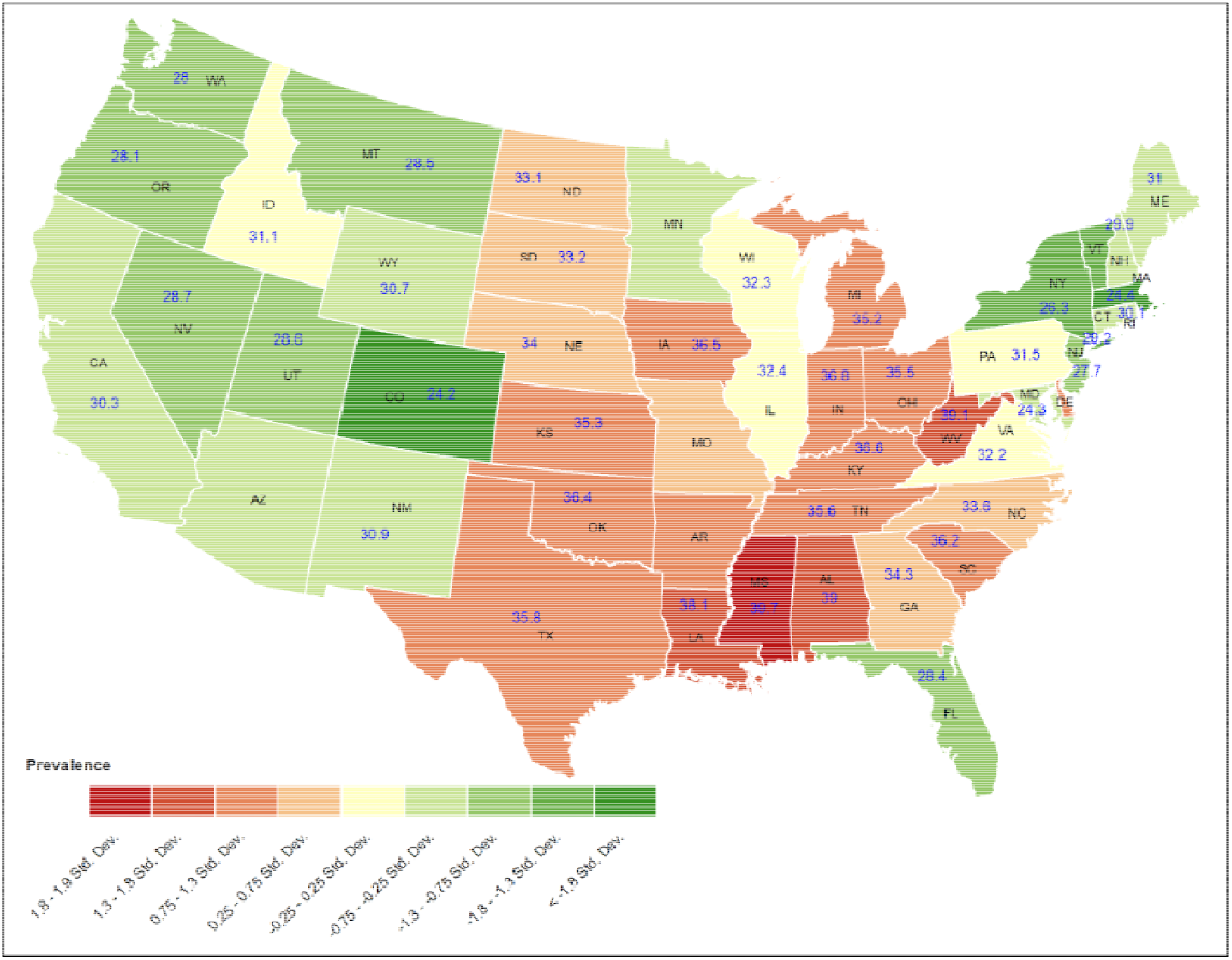
Overall adult obesity prevalence in the contiguous US in 2020. The number inside each state is the percent of prevalence. The half standard deviation map clearly shows the Data Source: The data comes from the Behavioral Risk Factor Surveillance System, a state-based telephone interview survey conducted by CDC and state health departments, BRFSS, 2020 (CDC, 2020b).

In the US, and even within the southern states, rural areas have been known to have a higher prevalence of obesity than urban areas, both for children and adults [5–9]. It could be noted that there is no universal agreement about which area can be defined as urban or rural, not even among the US federal agencies. Furthermore, it is impractical to designate an area as entirely rural or urban. In this context, the Economic Research Service (ERS) of the US Department of Agriculture (USDA) has developed different scales of rurality, such as Rural-Urban Continuum Codes (RUCC), Urban-Influence Codes (UIC), Rural-Urban Commuting Areas (RUCA), Frontier and Remote (FAR) Area Codes for a variety of geographic units (USDA, 2022a) [10]. The other sub-county-level urbanicity/rurality classification system, the RUCA codes, classifies US Census tracts based on population density, urbanization, and daily commuting measures (USDA, 2022b) [11].

Data with RUCA Codes are also available at the Zip Code level, which is apportionments from census tract data to the Zip Codes. Zip Code or its area-level equivalent Zip Code Tabulation Areas (ZCTA) is not the best choice for area-level public health study because of the approximation of secondary data interpolated from Census geographic units and due to changes in the Zip Code number or the area by the US Postal Service. However, when the health data are not available at any other sub-county level but only by Zip Code, it remains the only choice, and is still being widely used in the US for public health studies. As our health outcome data are available at the Zip Code level, we used RUCA codes in this study. Both Census tract-based and Zip Code-based RUCA have been used in many studies, often regrouping its 10-level classification into smaller groups [12-14]. Through a joint project, called PLACES, the CDC Foundation, in collaboration with the Robert Wood Johnson Foundation, provides health data for small areas. According to CDC, regardless of population size and rurality, these data allow us to better understand the burden and geographic distribution of health measures in their areas and assist them in planning public health interventions. One significant advantage of PLACES data is that they are available at different geographic units for the entire US. PLACES provides model-based health data to all counties, places (incorporated and census-designated places), Census tracts, and ZCTAs for major chronic disease measures. Many public health studies for different chronic diseases have been conducted using data from PLACES or its predecessor, the 500 Cities Project [15].

According to US Census Bureau, 19.3% of residents live in rural areas, where the healthcare infrastructure is not adequate, and there are barriers to accessing healthcare facilities [16-18]. Over the past decades, asthma and related allergic disorders (RAD) have increased in prevalence in the US [19], and as compared with rural areas, the prevalence of asthma in children and adults is higher in metropolitan and micropolitan areas [20]. However, another study shows asthma is more common in micropolitan areas than in rural and urban areas [20]. Additionally, diabetes and CVD are more prevalent among the rural population in the US [21]. Diabetes is another leading cause of death in the US and is reported to be up to 17% higher mortality in rural areas than in urban [21]. There has been evidence that diabetes is more prevalent in small towns and remote areas than in metropolitan areas, where according to Healthy People 2020, diabetes was ranked the third most significant healthcare priority in the local community [21]. Age-adjusted mortality for CVD and stroke was found to be higher in rural regions compared with urban or metropolitan areas [21-24]. Furthermore, rural patients suffer more stroke events but get minimal access to treatment during acute ischemic strokes than in urban areas [22]. Thus, according to the Reasons for Geographic and Racial Differences in Stroke study results, stroke risk was 23% higher in large agricultural towns, as well as 30% higher in small rural/isolated areas than it was in urban areas [23]. People in urban areas tend to have higher levels of cholesterol and low-density lipoprotein cholesterol than people in rural areas; therefore, residents of urban areas have less favorable lipid profiles as compared with residents of rural [24, 25]. In urban and rural public health, obesity causes many health problems both independently and together with other diseases. National surveys in the United States have shown a marked increase in the prevalence of obesity over time in the US, but obesity-related health outcomes are more common in the southern part of the US [26,27]. Several studies revealed that metropolitan residents suffer from high blood pressure problems more frequently than their rural counterparts [24, 30]

Figure 2 shows the distribution of the population according to RUCA codes grouped by southern states (MS, AL, LA, and GA,) and CDC measures of health outcomes per RUCA codes. The prevalence of obesity-related health outcomes increases from RUCA 1 to 10, but the total population decreases (Figure 2). Georgia, out of these four states, has the lowest rate of obesity, high cholesterol, CHD, and high blood pressure in its entire commuter area compared with LA, AL, and MS. MS, on the other hand, has the highest rate of obesity. COPD rates in MS are lower than those in LA and AL in all RUCAs Code, and lower than those in GA in RUCA 5-10 (micropolitan, small towns, or rural areas). In comparison to LA, AL, and GA, MS has a lower CHD rate in all RUCAs; however, compared with the state of Georgia, MS has a higher rate except for RUCA 9-10 (rural areas). According to the CDC data, the rural areas (RUCA 9-10 of MS) have the lowest CHD rates of the southern states.

**Figure 2:**
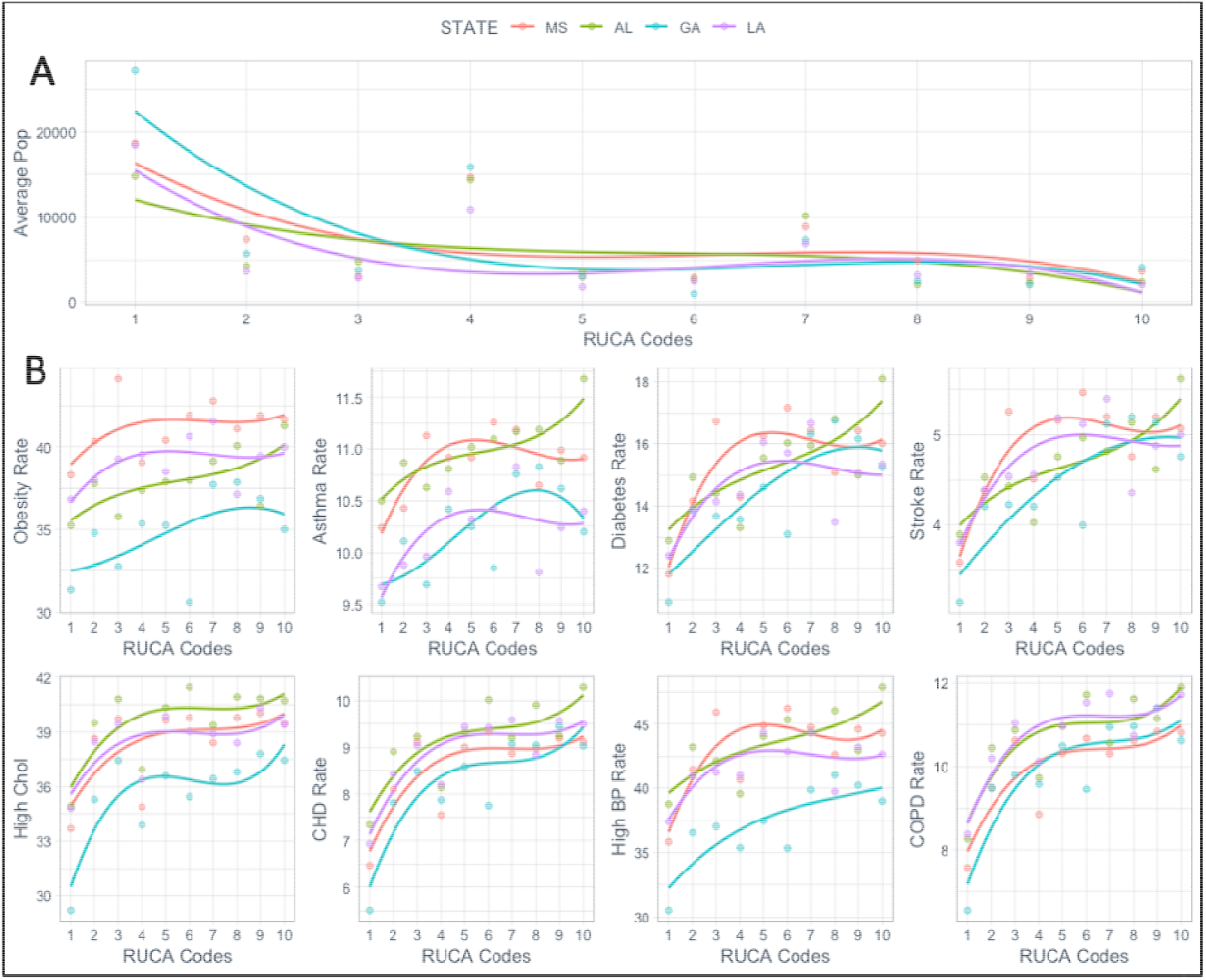
Lowess Smooth Curve showing the relationships of population and eight selected health outcomes with RUCA in MS, AL, GA, and LA. 2A). Proportion populations vary per RUCA codes; Georgia has the highest, and Alabama has the lowest urban population. As rurality increases, the population decreases in all four states. 2B). The patterns of health outcomes vary for all four states per the RUCA codes. For example, obesity rates for Georgia are the lowest and the highest for Mississippi across the RUCA codes. However, for most other outcomes, the rates are not the same across the RUCA codes.

Table 1 indicates that a total of 4,657,757 people reside in LA, 5,024,279 in AL, 2,961,279 in MS, and 10,711,908 in GA, with over half of them white. The majority of the population has high school diplomas, but only around 25% have a bachelor’s degree or higher (Table 1). Around 15% of the population under the age of 65 lives without health insurance in MS and GA, whereas in LA and AL, the percentage is a little bit lower. In the study area, the median household income in MS is the lowest, while the largest household income is in GA. In MS, over 19% of the population lives in poverty, which was the highest among these states, whereas Georgia had the lowest poverty rate (Table 1).

**Table 1.** Basic population characteristics of the study area.

| 2020 Data | Louisiana | Alabama | Mississippi | Georgia |
| --- | --- | --- | --- | --- |
| Population | 4,657,757 | 5,024,279 | 2,961,279 | 10,711,908 |
| White alone | 57.10% | 64.10% | 56.00% | 51.90% |
| Black alone | 31.40% | 25.80% | 36.60% | 31.00% |
| Hispanic or Latino | 6.90% | 5.30% | 3.60% | 10.50% |
| Asian alone | 1.90% | 1.50% | 1.10% | 6.00% |
| American Indian and Alaska Native alone | 0.70% | 0.70% | 0.60% | 0.50% |
| Diversity Index | 58.60% | 53.10% | 55.90% | 64.10% |
| High school graduate or higher, age 25+, 2016-2020 | 85.90% | 86.90% | 85.30% | 87.90% |
| Bachelor's degree or higher, age 25+, 2016-2020 | 24.90% | 26.20% | 22.80% | 32.20% |
| Persons without health insurance, underage 65 | 10.50% | 11.70% | 15.40% | 15.50% |
| Median household income, 2016-2020 | \$50,800 | \$52,035 | \$46,511 | \$61,224 |
| Per capita income in past 12 months, 2016-2020 | \$29,522 | \$28,934 | \$25,444 | \$32,427 |
| Persons in poverty | 17.80% | 14.90% | 18.70% | 14.00% |

## 2. Materials and Methods

### Study Area

There were four states included in this study that were MS, GA, LA, and AL in the United States. These states have a unique culture as they belong to a cultural and geographic subregion in the Southern US, known as the Deep South. Among these four states, for health outcomes, MS, LA, AL, and GA have been ranked at the bottom in the US.

### Data Sources

We used publicly available Zip Code level approximations of USDA-developed Rural-Urban Commuting Areas (RUCA) for rurality designation and Zip Code level PLACES data developed by CDC for health outcomes designation. The RUCA codes and descriptions are shown in Table 2. The selected obesity-related health outcomes were Asthma, COPD, Diabetes, High Cholesterol, High Blood Pressure, Coronary Heart Disease and Stroke. We used all categories of this 10-tired classification scheme, and for modeling, but for more strength of inference, we regrouped into three categories. For health outcomes, we used Zip Code Tabulation Areas (ZCTAs)-based PLACES data available from CDC’s website. We linked PLACES data and RUCA data using Zip Codes.

**Table 2.** Primary RUCA codes, 2010.

| Code | Classification description |
| --- | --- |
| 1 | **Metropolitan core:** primary flow within an urbanized area (UA) |
| 2 | **Metropolitan high commuting:** primary flow 30% or more to a UA |
| 3 | **Metropolitan low commuting:** primary flow 10% to 30% to a UA |
| 4 | **Micropolitan core:** primary flow within an urban cluster of 10,000 to 49,999 (large UC) |
| 5 | **Micropolitan high commuting:** primary flow 30% or more to a large UC |
| 6 | **Micropolitan low commuting:** primary flow 10% to 30% to a large UC |
| 7 | **Small town core:** primary flow within an urban cluster of 2,500 to 9,999 (small UC) |
| 8 | **Small town high commuting:** primary flow 30% or more to a small UC |
| 9 | **Small town low commuting:** primary flow 10% to 30% to a small UC |
| 10 | **Rural areas:** primary flow to a tract outside a UA or UC |
| 99 | **Not coded:** Census tract has zero population and no rural-urban identifier information |

### Partial Least Square Discriminant Analysis (PLS-DA) Methods

The PLS-DA method is utilized for the separation of samples into groups based on two data matrices, X (health outcomes) and Y (RUCAs). This method allows for the analysis of multiple categorical dependent variables simultaneously, and a linear subspace of explanatory variables is found to maximize the correlation between independent variables and corresponding dependent variables [28, 29]. By reducing the number of factors, this new subspace enables a simplified classification of dependent variables. A Variable Important Projection (VIP) score reflects a variable’s importance in the PLS-DA model and describes how it contributes to the model. The VIP score of a variable is defined as the sum of the square correlations between the PLS-DA components and the original variable, where the square correlation represents the amount of variation explained by the PLS-DA component in the model [29]. The number of terms in the sum depends on the number of PLS-DA components found to be significant in identifying the classes [30]. VIP scores are indicated on the Y axis, corresponding to the variables on the X axis. VIP scores provide the most contributing variables to class discrimination in the PLS-DA model. According to the results of this analysis, selected standardized health outcomes were associated with the RUCA Code. The VIP score is the statistical indicator of the significance of these relationships. When the VIP scores of selected health outcomes from the PLS-DA are greater than or equal to 1, then these health outcomes are considered to be significantly associated with the corresponding RUCA Code [31].

### Random Forest (RF) Method

As with the PLS-DA approach, the RF approach is very similar and is used for classifying and evaluating the most significant contributory variables in class discrimination. It can be described as a supervised nonlinear representation of class-level classification for categorical outcomes [32]. RF method provides the most important features, which are strongly associated with the categories outcomes such as RUCA [32]. Through random forest analysis, supervised learning algorithms with minimal errors can be incorporated to improve decision tree performance [33]. In this analysis, the random forest is fundamental, and it shows, each variable, how important it is in classifying the data [33]. The Mean Decrease Accuracy plot shows how much accuracy the model loses when each variable is excluded. The greater the drop-off inaccuracy, the more important the variable is for classification. In order of importance, the variables are listed in ascending order. The Gini coefficient is used as a measure of homogeneity in the resulting random forest. The study considered selected standardized health outcomes to be input features and RUCAs as output variables. By using Variables Importance Plots, we visualized the relationship between health outcomes and the RUCA Code. Similar results and inferences can be made in PLS-DA modeling approach. Using the mean decreased accuracy score, we were able to identify which health outcomes were strongly associated with which RUCA code.

### Multinomial logistic regression

The statistical correlation method is generally used to evaluate relationships for binary ouctcomes; however, for more than two groups of outcomes, multinomial logistic regression is meaningful for classification. The ten-tire RUCA classes have been grouped into three categories-a) RUCA 1-3: Metropolitan Area, b) RUCA 4-6: Micropolitan Area, and c) RUCA 7-10: Small towns or rural areas as done in a previous study [34-36]. We compared Metropolitan and Micropolitan areas with small towns or rural areas. In terms of independent variables, the health outcomes of the CDC were treated as independent variables, whereas the RUCA categories were considered as grouped outcomes. In general, a multinomial logistic model can be considered a logistic model for multiple dependent outcomes. Taking category J as the baseline category, then the logit for the baseline category is as follows:

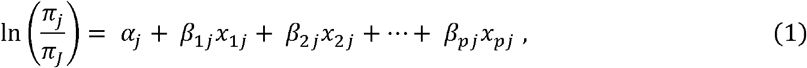

Where, j = 1, 2, …, J − 1 and is the number of predictors; *π*_*j*_ is the probability of each category of the dependent variables *α*_*j*_ denotes the intercept; β’s denotes the regression coefficient of the independent *variables x*. According to our data, the multinomial regression model will be

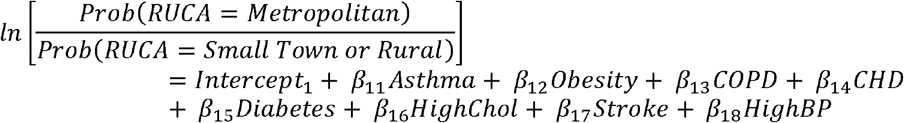

Similarly, other output will be

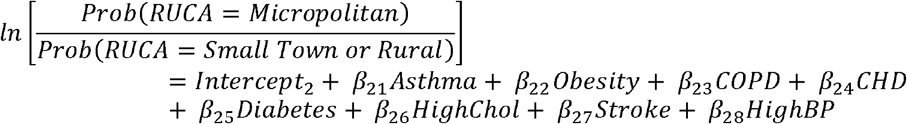

We considered small towns/rural areas as a baseline category, and we evaluated the effect of health outcomes associated with metropolitan and micropolitan areas. The full factorial model in statistical software R was employed to analyze the regression coefficients.

## 3. Results

As illustrated in Figure 4, health outcomes are clustered into two major groups (Cluster-1 and Cluster-2), in which each outcome is highly clustered with the other selected health outcomes. Cluster-1 showed an enriched positive association among obesity, asthma, stroke, diabetes, and high cholesterol in all RUCAs codes except 1 and 4 (Figure 4). Additionally, CHD, COPD, and high cholesterol are clustered in all RUCAs except RUCA 1, 2, and 4, as shown in Cluster-2. According to Figure 4 except for RUCA 1, RUCA 2, and RUCA 3, in all other areas, obesity, asthma, and stroke coexist with each other. In the rural areas (RUCA 6 - 10), most of the health outcomes are coexisting, indicating a higher disease burden in these areas.

According to our multivariate analysis results, COPD is significantly associated with RUCA 2, 7 and 10, while it is less associated with RUCA 1, 3, and 5 (VIP ≥ 1.0). Cardiovascular disease and high cholesterol are also highly associated with the RUCA 2, 7, and 10 areas (Figure 5A) and show lower association with the RUCA 1, 3, and 5 areas (Figure 5B). For RUCA 7 and 10, high cholesterol is a significantly highly associated health outcome; however, in RUCA 1 and 5, high cholesterol is a significantly less correlated health outcome. The random forest model yielded approximately the same results for finding the most important health outcome corresponding to the RUCA (Figure 5B). According to mean decreased accuracy and VIP score, COPD is the most significant prevalence that is strongly associated with RUCA 2 and 7-10. Based on the statistical method PLS-DA and the machine learning model random forest, we came up with nearly the same results for other health outcomes.

Figure 3A and Figure 3B show the crude (unadjusted) and standardized outcomes for asthma, obesity, COPD, CHD, diabetes, high cholesterol, high blood pressure, and strokes. The lowess smooth curve, shown in Figure 2, illustrates selected health outcomes by rural-urban commuting area codes. All the rural-urban commuter areas found MS to have the highest obesity rate in comparison with LA, GA, and AL, while GA had the lowest (Figure 2). Further, MS has lower CHD rates in all RUCAs compared with LA and AL, but compared with GA, MS has higher CHD rates except for RUCA 9-10 (rural areas). Consequently, the rural areas of Mississippi (RUCA 9-10) have the lowest CHD rates in the study area.

**Figure 3:**
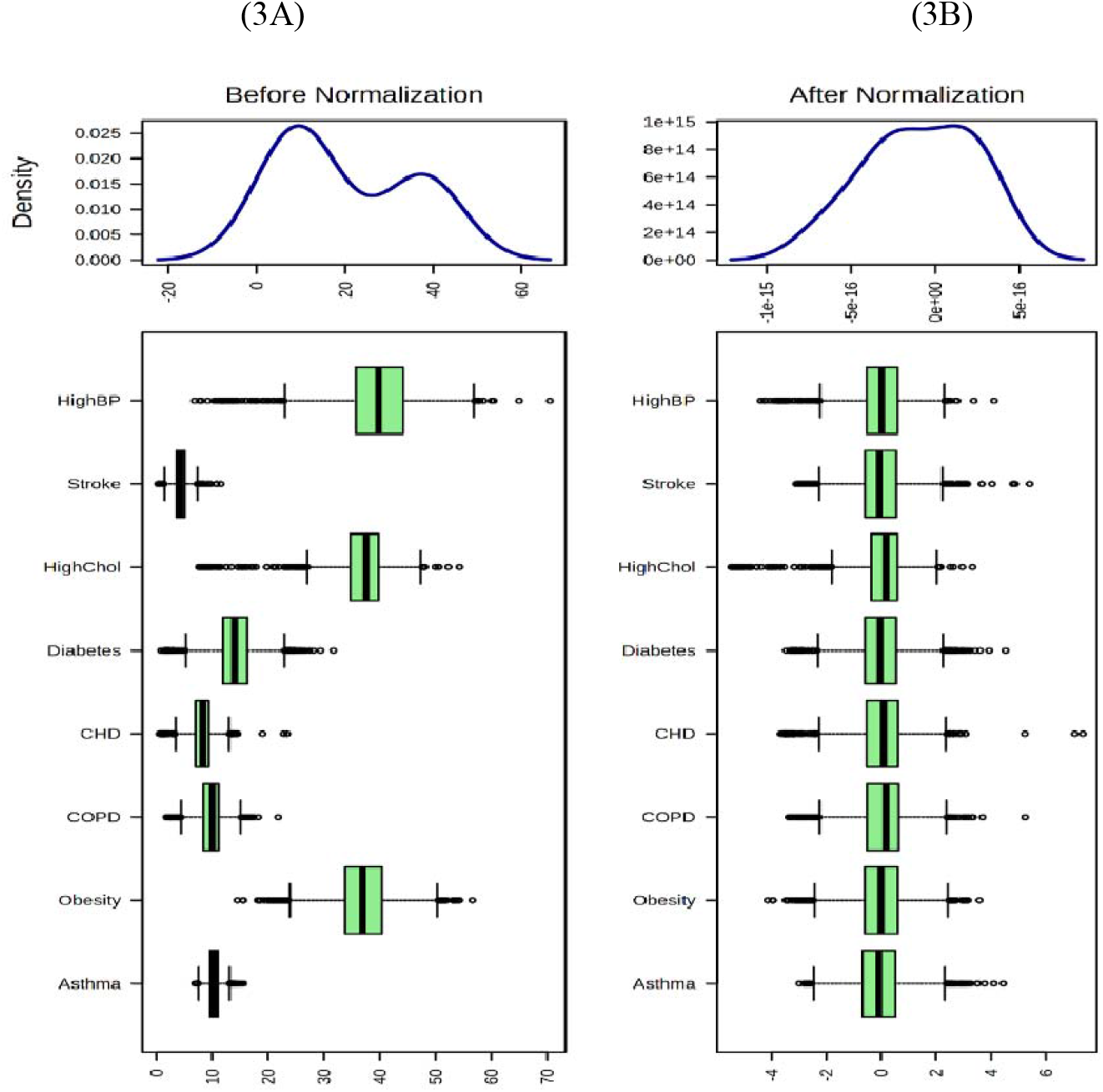
Multivariate statistical analysis for studying the associations of health outcomes with RUCAs. (A) Overall average rate of the eight health outcomes in the study area, (B) Standardized rates of the health outcomes.

**Figure 4:**
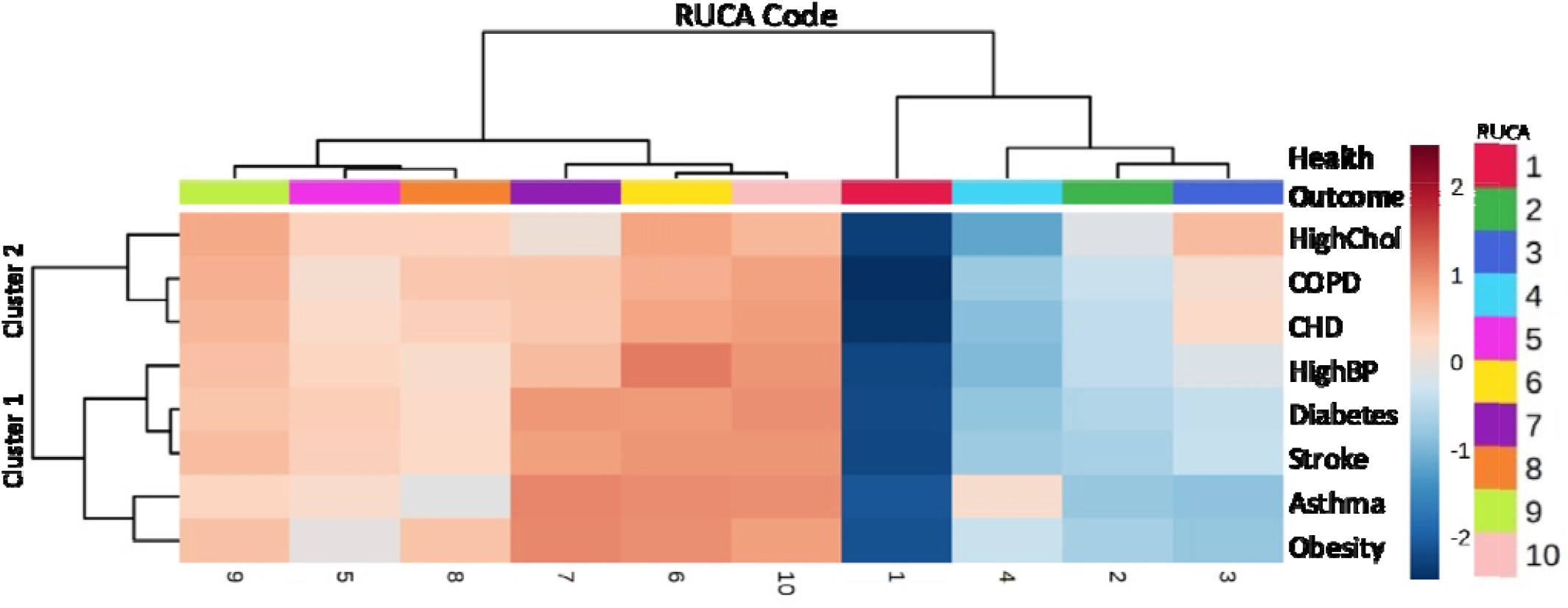
Hierarchical clustering of the health outcomes in association with RUCAs. Here clustering is demonstrating the magnitude of disease burden due to multiple health outcomes per RUCA.

**Figure 5:**
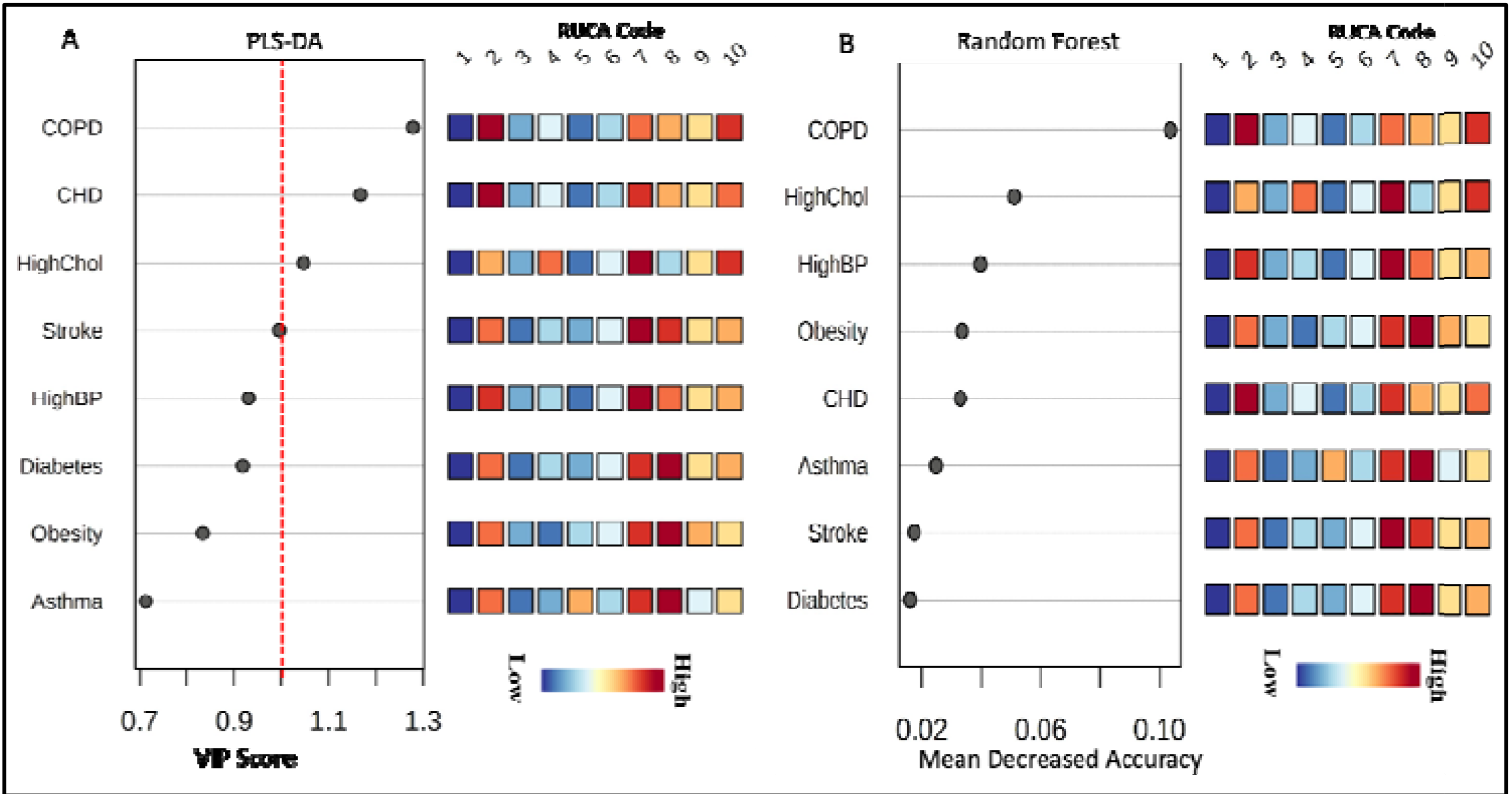
Multivariate statistical analysis for studying health outcomes and RUCA. (A) Variable Importance Projection according to RUCA by PLS-DA, (B) Variable Importance (VI) according to RUCA by Random Forest method.

As a means of gaining more strength of evidence, we regrouped the RUCAs into three major categories: Metropolitan (RUCA 1-3), Micropolitan (RUCA 4-6), and Small Town or Complete Rural (RUCA-7-10). A multinomial logistic regression model was used to assess the association between rural-urban commuting areas and selected health outcomes. Table 3 presents odds ratios for metropolitan and micropolitan areas with small towns or rural areas. A significant association between asthma prevalence and urban areas was observed. A 35% higher prevalence of asthma was found in micropolitan areas compared to small towns or rural areas (OR: 1.35, p = 0.001) when other health outcomes were held constant. Nonetheless, in metropolitan areas and micropolitan areas, obesity problems were significantly lower (46% and 27%, respectively) than in rural and small towns (OR: 0.54, p<0.001, OR: 0.63, p<0.001) respectively. Compared to both metropolitan (OR: 0.28, p<0.001) and micropolitan (OR: 0.27, p<0.001) areas, small towns or rural areas with COPD have significantly higher prevalence health outcomes. According to our Multinomial Logistic Regression Model, there is no significant association between CHD with any of the three categories in this study area.

**Table 3:**
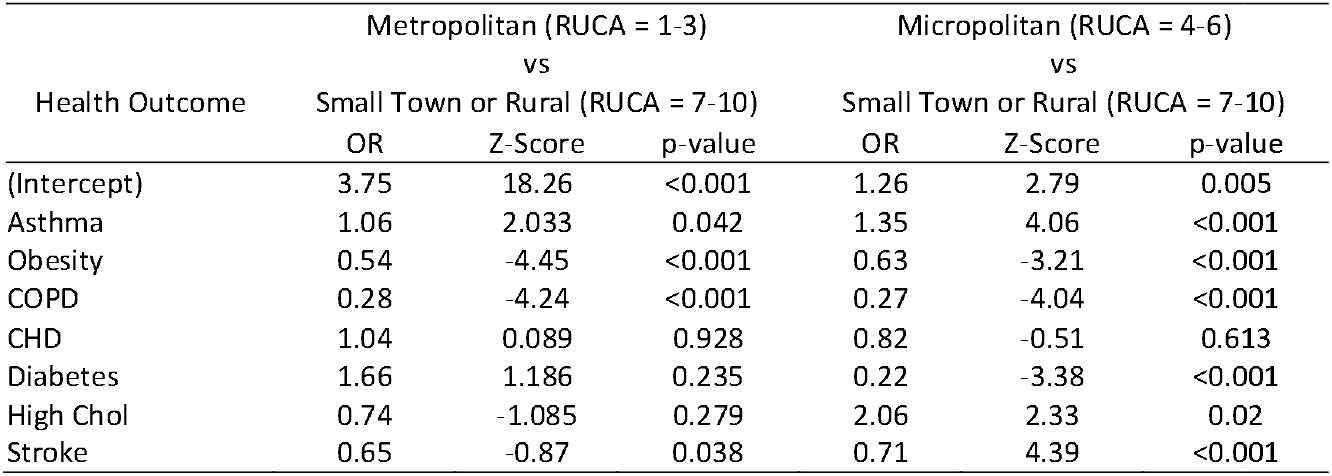
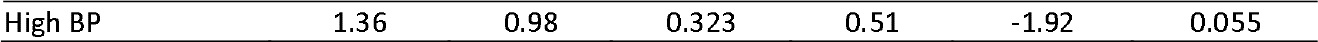
Multinomial Logistic Regression Model outcomes for Metropolitan vs. Rural and Micropolitan vs. Rural with selected Health outcome.

According to Table 3, there is a significant difference in the prevalence of Asthma, Obesity, COPD, and Stroke between Metropolitan and small towns or complete rural areas. However, CHD, Diabetes, High Cholesterol, and High BP do not show any significant difference between these two categories. On the other hand, while considering Micropolitan and small towns or complete rural areas, Asthma, Obesity, COPD, Diabetes, High Cholesterol, and Stroke show significant differences in prevalence. Only CHD and High BP do not have significant differences between these two categories.

## 4. Discussion

This study extended by analyzing commuting rurality (RUCA) in the southern part of the US. The main focus of the study was the major health problems associated with obesity-related health outcomes in rural trends in the southern parts of the United States. Despite substantial improvements in health and well-being for all Americans, health disparities between geographic and population groups have persisted and remained a significant concern for public health policy. As mentioned earlier, there are a variety of health outcomes, such as obesity, asthma, COPD, CHD, stroke, diabetes, high cholesterol, and high blood pressure showing striking disparities across the US [38]. This study attempts to reveal such disparities per the degree of urbanicity or rurality in the study area.

Studies show that asthma prevalence rates for children and adults are significantly higher in suburban and micropolitan areas than in rural areas [35, 36]. People living in urban areas are at a higher risk of asthma-related health outcomes than people living in rural areas [38]. Asthma is a significant health issue that affects one’s wellness for the rest of his or her life, taking into account the types of environments in which they live [38,39]. In this study, we found that the prevalence of asthma was associated with metropolitan (RUCA 1-3) and micropolitan areas (RUCA 4-6). Therefore, we conclude, consistent with other studies, asthma prevalence is higher in metro and micropolitan areas than it is in small towns or rural areas in the southern part of the United States.

We found a significant association of obesity prevalence with the spatial commuting pattern. Our research indicates that obesity is more prevalent in rural areas (RUCA 7-10), which is consistent with other studies [41]. Existing studies indicate that the prevalence of COPD in rural areas is higher than those in urban areas [42,43]. According to our study, among the selected health outcomes, COPD is the most significant health problem in RUCA 2 and RUCA 10. This study also shows that the prevalence of COPD in LA is higher than that in MS, AL, and GA in all RUCAs. Therefore, we gather that people living in smaller towns and rural areas suffer from higher rates of COPD than people living in major cities or micropolitan areas.

Diabetes is one of the leading causes of death and disability in rural areas of the United States [44]. Additionally, the prevalence of diabetes in rural areas is significantly higher by 17% than in urban areas [45]. Our study is aligned with these studies revealing a significantly lower prevalence of diabetes in micropolitan areas than in rural or small towns. However, we did not find any evidence of a significant difference in the prevalence of diabetes between metropolitan areas and small towns, but diabetes prevalence was strongly associated with RUCA 7 and 8.

There were more acute ischemic strokes among rural residents, and they had fewer treatment options. According to national data, rural populations are more likely to suffer strokes than urban populations in the United States. Our study results are consistent with the previous studies. Per our study, persons living in metropolitan areas have a 35% lower prevalence of stroke compared with those who live in rural areas or small towns. Furthermore, those who live in micropolitan areas (RUCA 4-6) have a 29% lower prevalence of stroke compared to those living in rural or small towns (RUCA 7-10). Consistent with the previous studies [45, 46], we find that stroke rates in the south are significantly higher in rural areas (RUCA 7-10).

The prevalence of high blood pressure and high cholesterol appears to be higher in urban areas compared with rural areas in the US [46], which is consistent with our findings. The high cholesterol rate and CHD rate in AL are higher in all RUCAs, compared to MS, GA, and LA. On the other hand, GA has a lower rate of high cholesterol compared with MS, AL, and LA. There is no significant difference in the prevalence of high cholesterol among metropolitan and small towns or rural areas in our study area. However, the prevalence of high cholesterol is almost twice in micropolitan areas in comparison with small towns or rural areas.

## 5. Conclusions

Using publicly available data, we utilized standardized rates from the CDC for selected health outcomes and the USDA RUCA code for the degree of rurality. In our study area, the prevalence of obesity, COPD, and stroke is greater in rural areas (RUCA7-10) than in urban areas, while the prevalence of asthma is higher in urban areas (RUCA1-3). Among the selected obesity-related health outcomes, COPD is the most significant and has a strong association with RUCAs 2, 7, and 10, whereas RUCAs 1 and 5 have a less significant relationship. These findings can be useful to target specific geographic areas for interventions.

## Data Availability

USDA, ZIP5. all the data are publicly available online

https://data.nal.usda.gov/dataset/pesticide-data-program-pdp/resource/10cf1c4d-57f5-4147-9187-44ce3ee683e2

## References

1. Petersen R, Pan L, Blanck HM. Racial and Ethnic Disparities in Adult Obesity in the United States: CDC’s Tracking to Inform State and Local Action. Prev Chronic Dis. 2019;16:E46.

2. CDC. Overweight & Obesity, Why it matters. https://www.cdc.gov/obesity/about-obesity/why-it-matters.html, accessed August 27, 2022.

3. Ward ZJ, Bleich SN, Cradock AL, Barrett JL, Giles CM, Flax C, et al. Projected U.S. State-Level Prevalence of Adult Obesity and Severe Obesity. N Engl J Med. 2019;381(25):2440–50.

4. CDC. Overweight & Obesity, Data & Statistics. https://www.cdc.gov/obesity/data/index.html. Accessed August 27, 2022.

5. Patterson PD, Moore CG, Probst JC, Shinogle JA. Obesity and physical inactivity in rural America. The Journal of Rural Health. 2004 Mar;20(2):151–9.

6. Lundeen EA, Park S, Pan L, O’Toole T, Matthews K, Blanck HM. Obesity prevalence among adults living in metropolitan and nonmetropolitan counties—United States, 2016. Morbidity and Mortality Weekly Report. 2018 Jun 6;67(23):653.

7. Trivedi T, Liu J, Probst J, Merchant A, Jones S, Martin AB. Obesity and obesity-related behaviors among rural and urban adults in the USA. Rural and remote health. 2015 Dec 1;15(4):217–27.

8. Johnson III JA, Johnson AM. Urban-rural differences in childhood and adolescent obesity in the United States: a systematic review and meta-analysis. Childhood obesity. 2015 Jun 1;11(3):233–41.

9. Voss JD, Masuoka P, Webber BJ, Scher AI, Atkinson RL. Association of elevation, urbanization and ambient temperature with obesity prevalence in the United States. International journal of obesity. 2013 Oct;37(10):1407–12.

10. USDA. Rural Classifications. Last updated June 17, 2021; Available online: https://www.ers.usda.gov/topics/rural-economy-population/rural-classifications/ (accessed on 6 September 2022).

11. USDA. 2010 Rural-Urban Commuting Area (RUCA) Codes. 2019. Available online: https://www.ers.usda.gov/data-products/rural-urban-commuting-area-codes/documentation (accessed on 6 September 2022).

12. Pruitt SL, Eberth JM, Morris ES, Grinsfelder DB, Cuate EL. Rural-urban differences in late-stage breast cancer: do associations differ by rural-urban classification system?. Texas public health journal. 2015;67(2):19.

13. Onega T, Weiss JE, Alford□Teaster J, Goodrich M, Eliassen MS, Kim SJ. Concordance of rural□urban self□identity and zip code□derived rural□urban commuting area (RUCA) designation. The Journal of Rural Health. 2020 Mar;36(2):274–80.

14. Gallagher A, Liu J, Probst JC, Martin AB, Hall JW. Maternal obesity and gestational weight gain in rural versus urban dwelling women in South Carolina. The Journal of Rural Health. 2013 Jan;29(1):1–1.

15. Monaghan A, Jones L, Brink L, Hacker K. Comparison of Census Tract–Level Chronic Disease Prevalence Estimates From 500 Cities and Local Health Claims Data. Journal of Public Health Management and Practice. 2022 Jan 1;28(1):E92–5.

16. Ratcliffe M, Burd C, Holder K, Fields A. Defining rural at the US Census Bureau. American community survey and geography brief. 2016 Dec 8;1(8):1–8.

17. Laditka JN, Laditka SB, Probst JC. Health care access in rural areas: evidence that hospitalization for ambulatory care-sensitive conditions in the United States may increase with the level of rurality. Health & place. 2009 Sep 1;15(3):761–70.

18. Okobi OE, Ajayi OO, Okobi TJ, Anaya IC, Fasehun OO, Diala CS, Evbayekha EO, Ajibowo AO, Olateju IV, Ekabua JJ, Nkongho MB. The Burden of Obesity in the Rural Adult Population of America. Cureus. 2021 Jun 20;13(6).

19. Martinez FD. Role of viral infections in the inception of asthma and allergies during childhood: could they be protective?. Thorax. 1994 Dec;49(12):1189.

20. Guo Z, Qin X, Pate CA, Zahran HS, Malilay J. Asthma Among Adults and Children by Urban–Rural Classification Scheme, United States, 2016-2018. Public Health Reports. 2021 Oct 4:00333549211047552.

21. O’Connor A, Wellenius G. Rural–urban disparities in the prevalence of diabetes and coronary heart disease. Public health. 2012 Oct 1;126(10):813–20.

22. Koifman J, Hall R, Li S, Stamplecoski M, Fang J, Saltman AP, Kapral MK. The association between rural residence and stroke care and outcomes. Journal of the neurological sciences. 2016 Apr 15;363:16–20.

23. Saad AM, Abushouk AI, Al-Husseini MJ, Salahia S, Alrefai A, Afifi AM, Abdel-Daim MM. Characteristics, survival and incidence rates and trends of primary cardiac malignancies in the United States. Cardiovascular Pathology. 2018 Mar 1;33:27–31.

24. de Groot R, van den Hurk K, Schoonmade LJ, de Kort WL, Brug J, Lakerveld J. Urban-rural differences in the association between blood lipids and characteristics of the built environment: a systematic review and meta-analysis. BMJ global health. 2019 Jan 1;4(1):e001017.

25. Cohen SA, Nash CC, Byrne EN, Mitchell LE, Greaney ML. Black/White Disparities in Obesity Widen with Increasing Rurality: Evidence from a National Survey. Health Equity. 2022 Mar 1;6(1):178–88.

26. Lutfiyya MN, Lipsky MS, Wisdom□Behounek J, Inpanbutr□Martinkus M. Is rural residency a risk factor for overweight and obesity for US children?. Obesity. 2007 Sep;15(9):2348–56.

27. Majumdar S, Morris A, Gordon C, Kermode JC, Forsythe A, Herrington B, Megason GC, Iyer R. Alarmingly high prevalence of obesity in haemophilia in the state of Mississippi. Haemophilia. 2010 May;16(3):455–9.

28. Mohamad N, Ismet RI, Rofiee M, Bannur Z, Hennessy T, Selvaraj M, Ahmad A, Nor F, Abdul Rahman T, Ismail A, Teh LK. Metabolomics and partial least square discriminant analysis to predict history of myocardial infarction of self-claimed healthy subjects: validity and feasibility for clinical practice. Journal of clinical bioinformatics. 2015 Dec;5(1):1–0.

29. Marzetti E, Landi F, Marini F, Cesari M, Buford TW, Manini TM, Onder G, Pahor M, Bernabei R, Leeuwenburgh C, Calvani R. Patterns of circulating inflammatory biomarkers in older persons with varying levels of physical performance: a partial least squares-discriminant analysis approach. Frontiers in medicine. 2014 Sep 1;1:27.

30. Zhang L, Zhang S, Sun M, Wang Z, Li H, Li Y, Li G, Lin L. Blood species identification using Near-Infrared diffuse transmitted spectra and PLS-DA method. Infrared Physics & Technology. 2016 May 1;76:587–91.

31. Oussama A, Elabadi F, Platikanov S, Kzaiber F, Tauler R. Detection of olive oil adulteration using FT-IR spectroscopy and PLS with variable importance of projection (VIP) scores. Journal of the American Oil Chemists’ Society. 2012 Oct;89(10):1807–12.

32. Mahmood A, Wang JL. Machine learning for high performance organic solar cells: current scenario and future prospects. Energy & environmental science. 2021;14(1):90–105.

33. Zimmerman N, Presto AA, Kumar SP, Gu J, Hauryliuk A, Robinson ES, Robinson AL, Subramanian R. A machine learning calibration model using random forests to improve sensor performance for lower-cost air quality monitoring. Atmospheric Measurement Techniques. 2018 Jan 15;11(1):291–313.

34. Nelson WA, Rosenberg MC, Mackenzie T, Weeks WB. The presence of ethics programs in critical access hospitals. InHEC forum 2010 Dec (Vol. 22, No. 4, pp. 267–274). Springer Netherlands.

35. French DD, Bradham DD, Campbell RR, Haggstrom DA, Myers LJ, Chumbler NR, Hagan MP. Factors associated with program utilization of radiation therapy treatment for VHA and medicare dually enrolled patients. Journal of community health. 2012 Aug;37(4):882–7.

36. Krstić G. Asthma prevalence associated with geographical latitude and regional insolation in the United States of America and Australia. PLoS One. 2011 Apr 8;6(4):e18492.

37. Hammond G, Luke AA, Elson L, Towfighi A, Joynt Maddox KE. Urban-rural inequities in acute stroke care and in-hospital mortality. Stroke. 2020 Jul;51(7):2131–8.

38. Wright RJ, Fisher EB. Putting asthma into context: community influences on risk, behavior, and intervention. Neighborhoods and health. 2003 Mar 20:233–64.

39. Sullivan K, Thakur N. Structural and social determinants of health in asthma in developed economies: a scoping review of literature published between 2014 and 2019. Current allergy and asthma reports. 2020 Feb;20(2):1–2.

40. Al-Kindi SG, Brook RD, Biswal S, Rajagopalan S. Environmental determinants of cardiovascular disease: lessons learned from air pollution. Nature Reviews Cardiology. 2020 Oct;17(10):656–72.

41. Cohen SA, Cook SK, Kelley L, Foutz JD, Sando TA. A closer look at rural□urban health disparities: associations between obesity and rurality vary by geospatial and sociodemographic factors. The Journal of Rural Health. 2017 Apr;33(2):167–79.

42. Panettieri Jr RA. Effects of corticosteroids on structural cells in asthma and chronic obstructive pulmonary disease. Proceedings of the American Thoracic Society. 2004 Nov;1(3):231–4.

43. Abrams TE, Vaughan-Sarrazin M, Fan VS, Kaboli PJ. Geographic isolation and the risk for chronic obstructive pulmonary disease–related mortality: A cohort study. Annals of internal medicine. 2011 Jul 19;155(2):80–6.

44. Glenn LE, Nichols M, Enriquez M, Jenkins C. Impact of a community□based approach to patient engagement in rural, low□income adults with type 2 diabetes. Public Health Nursing. 2020 Mar;37(2):178–87.

45. Harris JK, Beatty K, Leider JP, Knudson A, Anderson BL, Meit M. The double disparity facing rural local health departments. Annual review of public health. 2016 Mar 18;37:167–84.

46. Dodani S, Mistry R, Khwaja A, Farooqi M, Qureshi R, Kazmi K. Prevalence and awareness of risk factors and behaviours of coronary heart disease in an urban population of Karachi, the largest city of Pakistan: a community survey. Journal of public health. 2004 Sep 1;26(3):245–9.

